# Time In Range, as measured by continuous glucose monitor, as a predictor of microvascular complications in type 2 diabetes– A systemic review

**DOI:** 10.1101/2021.08.27.21262743

**Authors:** Rishi Raj, Rahul Mishra, Nivedita Jha, Vivek Joshi, Riccardo Correa, Philip A. Kern

## Abstract

**Aim:** Continuous glucose monitoring (CGM) derived times in range (TIR) correlates with hemoglobin A1c (A1c) among patients with type 2 diabetes mellitus (T2DM); however, there is a paucity of data evaluating its association with microvascular complications. We conducted this systematic review to examine the association between TIR and microvascular complications of diabetic retinopathy (DR), diabetic nephropathy (DN) and peripheral diabetic neuropathy (DPN).

**Method:** We conducted a comprehensive literature search on online database of PubMed, Scopus, and Web of Science following the Preferred Reporting Items for Systematic Reviews and Meta-Analyses (PRISMA) guidelines. Full texts original articles that evaluated association between CGM-derived TIR and risk of microvascular complications which were published between 2010 and June 2021, were included in our systematic review. The quality of included studies were evaluated using National Heart, Lung, and Blood Institute Quality Assessment Tool for Observational Cohort and Cross-Sectional Studies. Data were analyzed using qualitative synthesis.

**Result:** Eleven studies were included in the systematic review. The mean sample size, baseline A1c, and diabetes duration were 1271 (105-5901), 8.2 % (SD 0.5 %) and 11.3 years, respectively. Majority of studies were conducted in Asia (10 out 11). Four studies evaluated the relationship between CGM-derived TIR and DR and CGM-derived TIR and DN, while seven studies evaluated the relationship between CGM-derived TIR and DPN. A 10 % increase in TIR was associated with a reduction in albuminuria, severity of diabetic retinopathy, and prevalence of diabetic peripheral nephropathy and cardiac autonomic neuropathy. In addition, an association was observed between urinary albumin-to-creatinine ratio but not with estimated glomerular filtration rate.

**Conclusion:** This review summarizes recent evidence supporting an association between CGM-derived TIR and microvascular complications among patients with T2DM. A larger□scale multi-center investigation that includes more diverse participants is warranted to further validate the utility of TIR as a predictor for diabetic microvascular complications.

## INTRODUCTION

Hemoglobin A1c (A1c) is the primary tool for monitoring long-term glycemic control and assessing the risk of diabetes-related complications (1, 2). However, A1c has several limitations. A1c is affected by factors such as age, race/ethnicity, hemoglobinopathies, hemolytic anemia, recent blood transfusion, chronic kidney disease, and pregnancy, resulting in discrepancies between measured A1c and true glycemic control. Furthermore, A1c fails to provide information about extremes of hypo-or hyperglycemia, glucose trends, and glycemic variability (3). Intermittent self-monitored blood glucose (SMBG) is not influenced by conditions affecting red blood cell turnover and provides information beyond A1c; however, it is inconvenient and unpopular among patients of all age groups.

Continuous glucose monitoring (CGM) devices are increasingly popular, affordable, reliable in improving A1c, overcome many of the limitations with A1c and SMBG and CGM-derived metrics are now incorporated into the management of diabetic patients (4). The CGM devices measure interstitial fluid glucose every 1 to 5 minutes and provide 10 standardized core metrics, which include data on CGM use, mean glucose, time in range (70-180 mg/dL), and time above and below range. Time in Range (TIR), defined by the percentage of time glucose level is between 70 - 180 mg/dL (3.9-10.0 mmol/L), has been gaining popularity as a novel metric of glycemic control and strongly correlates with A1c in most studies (4). International consensus recommends TIR of 70% to align with A1c of ∼7%, with a 0.5% decline in A1c per 10% increase in TIR (4, 5). Furthermore, a 5 % increase in TIR was associated with significant clinical benefits among patients with type 2 diabetes mellitus (T2DM) (4).

While A1c remains the primary predictor for the development and progression of microvascular complications among patients with T2DM, there is growing evidence to support the association between TIR and diabetes-related microvascular complications (6-8). In a study by Sheng et al. on patients with T2D, authors calculated TIR from seven-point SMBG and found lower TIR to have higher odds of having diabetic retinopathy (DR), diabetic nephropathy (DN) and diabetic peripheral neuropathy (DPN), suggesting a strong correlation between calculated TIR and risk of diabetes-related microvascular complications (8).

Although adoption of TIR in clinical practice is gradually increasing and becoming well established, its use as a predictor for long-term risk of diabetes-related microvascular complications is still growing and needs further validation. Therefore, we performed a systematic review to summarize the published literature evaluating CGM-derived TIR as a predictor for diabetes-related microvascular complications, and discuss its implications on future clinical practice and research among patients with diabetes.

## METHODS

This systematic review was performed using the guidelines established by Preferred Reporting Items for Systematic Reviews and Meta-analyses (PRISMA) statements. (9) For this study, a comprehensive literature search was done to identify the original research articles in various databases. Before the literature screening process, a protocol was submitted to the International Prospective Register of Systematic Reviews (PROSPERO).

### Selection Criteria

Studies included in this systematic review met the following inclusion criteria: (a) cross-sectional/observational studies examining the association between CGM-derived TIR and microvascular complications among patients with T2DM (b) full-text and (c) English language articles (d) published between January 1, 2010, and June 5, 2021. We excluded the following studies: (a) review articles and (b) systematic review with or without meta-analysis.

### Search Strategy

On June 5, 2021, we conducted a comprehensive literature search in the electronic databases (PubMed, Scopus, and Web of Science) for publications between 2010 to June 2021 with English language restrictions. The search strategy was designed and conducted by the principal investigator (RR) with input from the study’s co-investigators (RM, NJ, and VJ) using the following keywords: ((“Continuous glucose monitor” OR “Continuous glucose monitoring” OR “Dexcom” “Freestyle Libre” OR “Guardian” OR “Flash glucose monitoring” OR “Time in Range” OR “Time-in-range” OR “TIR”) AND (“Diabetes complications” OR “Microvascular complications” OR “Retinopathy” “Neuropathy” OR “Nephropathy” OR “Complication” OR “Microalbuminuria” OR “Albuminuria”)) AND (“Type II diabetes” OR “type 2 diabetes”). Next, the bibliographies of the selected articles were manually searched for any additional studies. After screening for duplicate studies, two reviewers (RM and NJ) independently reviewed the title and abstract of the identified publications. Studies were then excluded if they did not address our research question, or meet our pre-specified inclusion criteria. Finally, the full texts of the remaining articles were examined to determine the final exclusion for our systematic review. Any conflicts during the study selection process were resolved by the third reviewer (RR). Figure 1 shows the schematic diagram of the study selection process.

**Figure 1.**
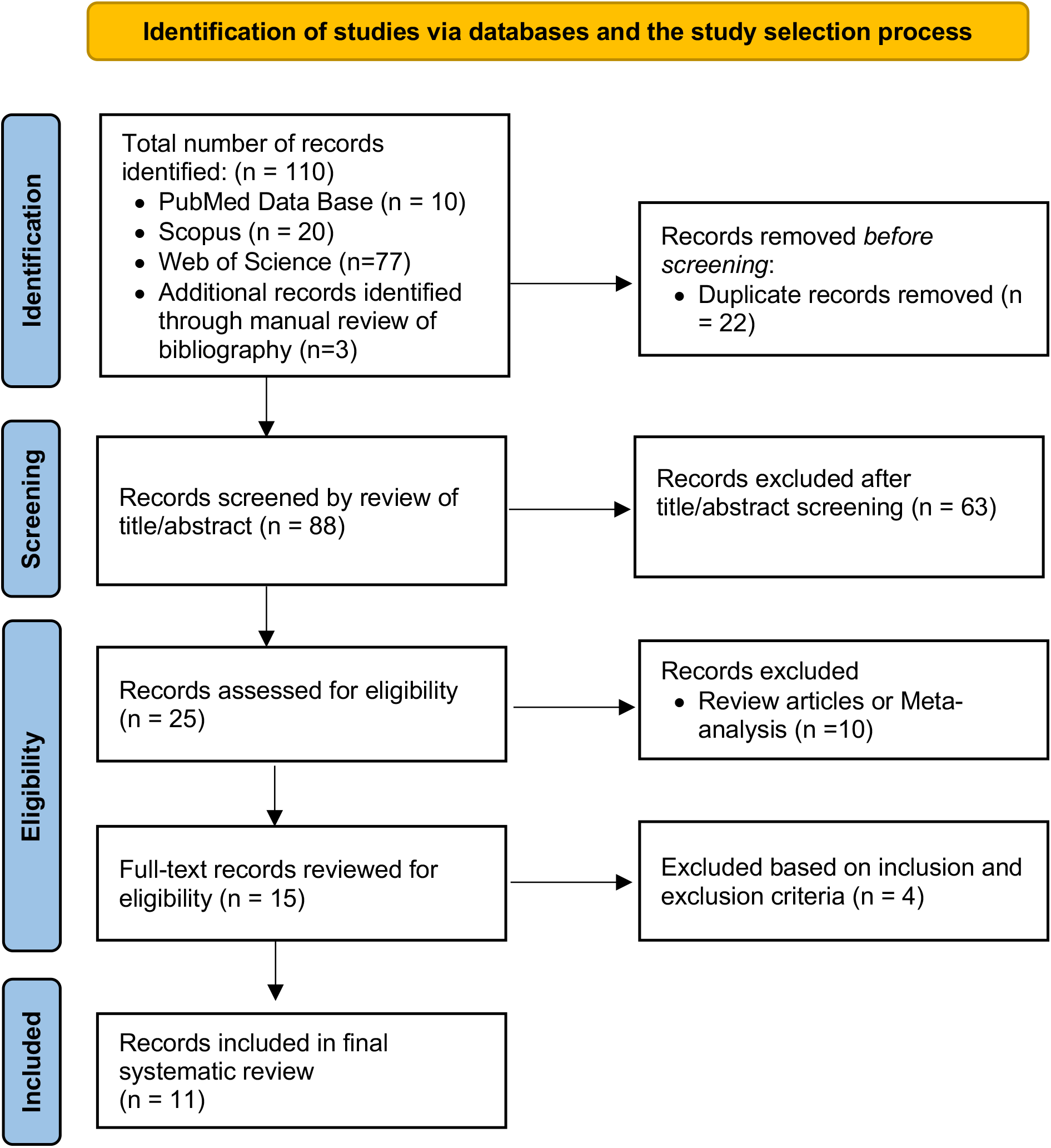
The PRISMA method flow diagram for study selection

### Data Collection

A predefined excel sheet was used for the extraction of data from each study. Following data were extracted from each included study: (a) title, (b) primary author, (c) year of publication, (d) duration of study, (e) country of the study population, (f) study design, (g) aim of the study, (h) sample size, (i) pertinent variables measure, (j) results, (k) conclusion, and (l) limitations of the study.

### Quality Assessment

Two independent reviewers (RM and VJ) assessed the study quality and risk of bias in the included studies using National Heart, Lung, and Blood Institute Quality Assessment Tool for Observational Cohort and Cross-Sectional Studies. (Study Quality Assessment Tools [https://www.nhlbi.nih.gov/health-topics/study-quality-assessment-tools] (10). Any disagreement was resolved by the other involved research reviewers (RR and VJ). Based on 14 questions, an overall rating was assigned as good, fair, and poor for study quality, indicating a corresponding low, moderate or high risk of bias.

### Data Synthesis

Before this systematic review, an initial literature search in the subject field revealed only a few studies with heterogeneous data points. Hence, we could not perform a meta-analysis, and a qualitative synthesis was performed.

## RESULTS

### Study Selection

We identified 110 publications using our initial search of online databases: PubMed, Scopus, Web of Science, and manual review of the bibliography. Of these, 22 duplicate publications were removed. Of 88 records screened, 63 were excluded based on screening of the title/abstract, and 10 were excluded being either review articles or meta-analysis. 15 eligible publications were retrieved in the full text, of which 4 did not meet inclusion criteria. Ultimately, we included 11 articles in our systematic review **(Figure 1)** (11-21). There was complete agreement between authors regarding article inclusion, risk-of-bias assessment, and data extraction. Detailed characteristics of included studies are summarized in **Supplementary File 1**.

### Baseline Characteristics

The mean sample size of included studies was 1271 and ranged from 105-5901 across 11 studies. Ten studies were cross-sectional in design, while one study was an interim analysis of an ongoing prospective observational study (19). The mean age of participants was 59.3 years (SD 1.3); A1c at baseline 8.2 % (SD 0.5 %); and duration of diabetes 11.3 years (SD 1.0). For patients with retinopathy, nephropathy, and neuropathy, the mean duration of diabetes was 11.8, 13.1, and 11.0 years, the mean age was 62.1, 61.6, and 59.1 years and the mean baseline A1c was 8, 7.8, and 8.1 %, respectively. Five studies were conducted in China (11-14, 20), two in South Korea (16, 17), two in Japan (19, 21), one in India (18) and one in the United States of America (15). Five studies used Medtronic’s CGM device (11, 14-17), four studies used Abbott’s Freestyle libre CGM (FLP-CGM) device (18-21), one study used Meiqi CGM device (13) while in one study both Medtronic and Meiqi CGM devices (12) were used. In all 11 studies, TIR was defined by the time percent during 24-hours when the glucose was in the range of 70-180 mg/dL (3.9-10 mmol/L). Duration of use of CGM device varied in all studies and ranged between 3 to 14 days. Calibrations of the CGM device, when applicable (in 6 of 11 studies) was done using two to four capillary blood glucose readings. In one study, the number of calibrations was not reported by the authors (14). Glucose management indicator values based on CGM data were available for only 5 studies and ranged between 7 to 7.5 (15-18, 20). We have summarized the baseline characteristics of the included studies in **Table 1**. On the study quality assessment tool, 1 study was rated “good”, and 10 studies were rated “fair”; and none were rated “poor”, eliminating increased risk of bias in the studies included in our systematic review **(Supplementary File 1)**.

**Table 1.** Baseline characteristics of the studies evaluating the association between CGM-derived TIR and microvascular complications among patients with T2DM.

| Characteristics | All included studies (n=11) | TIR and Diabetic Retinopathy (n=4) | TIR and Diabetic Nephropathy (n=4) | TIR and Diabetic Neuropathy (n=7) |
| --- | --- | --- | --- | --- |
| Sample size, n (mean) | 1271.5 | 370 | 2011.7 | 2703.2 |
| Sex (%) |  |  |  |  |
| Male | 60.8 | 58.1 | 62.0 | 62.6 |
| Female | 39.2 | 41.9 | 38.0 | 37.4 |
| Age, years (mean, SD)<sup>§</sup> | 59.3, 1.3 | 62.1, 0.99 | 61.6, 0.4 | 59.1, 2.8 |
| Baseline A1c, % (mean, SD)<sup>§</sup> | 8.2, 0.5 | 8.0, 0.6 | 7.8, 0.4 | 8.1, 0.6 |
| Duration of diabetes, years(mean, SD)<sup>†</sup> | 11.3, 1.0 | 11.8, 0.7 | 13.1, 0.2 | 11.0, 1.1 |
| Study location (n) |  |  |  |  |
| China | 5 |  |  |  |
| South Korea | 2 |  |  |  |
| Japan | 2 |  |  |  |
| India | 1 |  |  |  |
| United States of America | 1 |  |  |  |
| CGM device used (n) |  |  |  |  |
| Medtronic | 5 |  |  |  |
| Abbott Freestyle Libre | 4 |  |  |  |
| Meiqi | 1 |  |  |  |
| Medtronic + Meiqi | 1 |  |  |  |
| Duration of CGM use (n) |  |  |  |  |
| 3 days | 4 |  |  |  |
| 14 days | 4 |  |  |  |
| 3 or 6 days for **GOLD™** (Medtronic) and **iPro™2** (Medtronic), respectively* | 2 |  |  |  |
| Two 6-day periods, separated by 2 weeks<sup>#</sup> | 1 |  |  |  |
| CGM device calibrations (n) |  |  |  |  |
| Not applicable | 4 |  |  |  |
| At least 2 times per day | 3 |  |  |  |
| At least 4 times per day | 3 |  |  |  |
| Not reported | 1 |  |  |  |
| <sup>§</sup>Two studies (Yang et al. and Kuroda et al.) reported median age and median baseline A1c in their study and hence were not included in the final calculation of mean age and average baseline A1c of participants.<br><sup>†</sup>Four studies (Yang et al., Kuroda et al., Guo et al., and Guo et al.) reported median values for the duration of diabetes and hence were not included in the calculation of the mean duration of diabetes.<br>*Yoo et al. and Kim et al. used **GOLD™** (Medtronic) and **iPro™2** (Medtronic) CGM over 3 and 6 days, respectively.<br><sup>#</sup>Mayeda et al. collected CGM data over two 6-days periods, separated by 2 weeks. |  |  |  |  |

#### 1. Association between CGM-derived TIR and diabetic retinopathy (DR)

Four cross-sectional studies evaluated the association between TIR and DR (11, 18, 19, 21). One study used Medtronic CGM over 72 hours (11) while three studies used the FLP-CGM device to collect 14 days of data (18, 19, 21). One study used FLP-CGM data over the middle 8-day period, excluding the first 2 days and last 4 days (19). The average prevalence of DR was 27.4 % (range: 22.2 to 30.1%). One study categorized participants into three groups based on the glycemic profile (“TIR Profile”, “Hypo Profile”, and “Hyper Profile”) and showed higher odds of proliferative DR (“Hypo profile”: OR= 2.84, 95% CI: 1.65–4.88 and “Hyper profile”: OR =1.39, 95% CI: 0.78–2.45) as well as non-proliferative DR (“Hypo profile’’: OR =1.44, 95 % CI =1.20-1.73 and “Hyper profile’’: OR =1.33, 95 % CI = 1.11-1.58) compared to “TIR profile” (18). Two studies found a statistically significant association between a 10% increase in TIR and reduction in the severity of DR (11, 19) while no significant association between TIR and DR was found in the study by Kuroda et al. (21). In both these studies, higher A1c was found to be associated with increased severity of DR (p<0.01) (11, 19) while the relationship between A1c and DR was not assessed in the other two studies (18, 21).

In summary, a 10 % increase in TIR is associated with reduction in severity of DR and higher time spent in target range is associated with decrease in severity of DR. In addition, CGM-derived TIR was found to be similar to A1c in predicting DR among patients with T2DM.

#### 2. Association between CGM-derived TIR and diabetic nephropathy (DN)

The association between TIR and nephropathy was evaluated in four studies (16, 18, 19, 21). In one study, continuous glucose monitoring was done using Medtronic CGM device for 3 days (GOLD™) and 6 days (iPro^TM2^), respectively (16) while in three studies, FLP-CGM device was used to record data over 14 days (18, 19, 21). In three studies, urinary albumin-to-creatinine ratio (UACR) > 30 was used to define DN (16, 18, 19) while in one study, authors used UACR and EGFR values to determine its association with TIR (21). Participants were categorized into normoalbuminuria (UACR <30 mg/g), microalbuminuria (UACR 30-300 mg/g) or macroalbuminuria (UACR >300 mg/g) in two studies (18, 19). In one study, participants were grouped into with-and without-albuminuria (16). The prevalence of albuminuria in these four studies was 32.67 % (range 27-36.6 %), with a higher prevalence of microalbuminuria compared to macroalbuminuria (22.6 % vs. 8.5 %, respectively). In another study, participants were categorized into three groups based on the glycemic profile (“TIR Profile”, “Hypo Profile”, and “Hyper Profile”) (18). Compared to “TIR profile”, both “hyper” and “hypo” profiles had higher odds of macroalbuminuria (“Hypo profile”: OR= 1.58, 95% CI: 1.25–1.98; “Hyper profile”: OR = 1.37, 95% CI: 1.10–1.71). Additionally, “hyper” and “hypo” profiles also had higher odds of diabetic kidney disease, compared to “TIR profile” (“Hypo profile”: OR=1.65, 95% CI: 1.18– 2.31; “Hyper profile”: OR = 1.88, 95% CI: 1.37–2.58) (18). Two studies showed a statistically significant association between a 10% increase in TIR and a reduction in the severity of albuminuria (16, 19). In multiple regression analysis, Kuroda et al. found an association between TIR and UACR (β = −0.100, *P* = 0.043) but not with eGFR (β = −0.011, *P* = 0.824) (21). Out of the four studies evaluating relationship between TIR and DN, two studies found similar association between A1c and DN (16, 19). One study found higher A1c among patients with albuminuria compared to patient without albuminuria (8.5% vs 8.0%, p <0.01) (16) while in the second study, authors found a statistically significant association between severity of albuminuria and A1c (p <0.01) (19). In the rest of the two studies, the association between A1C and DN was not evaluated (18, 21).

In summary, two studies showed decrease in severity of albuminuria with a 10 % increase in TIR, one study showed increased time spent in target range to be associated with lower risk of macroalbuminuria and diabetic kidney disease, while in one study, increased TIR was associated with decreased in UACR but not with eGFR. In limited studies evaluating relationship between CGM-derived TIR to DN, it was found to be similar to A1c in predicting DN among patients with T2DM.

#### 3. Association between CGM-derived TIR and diabetic neuropathy

Seven studies examined the relationship between CGM-derived TIR and diabetic neuropathy (12-15, 17, 20, 21). Four studies evaluated the association of TIR with diabetic peripheral neuropathy (DPN) (13, 15, 20, 21), two studies examined the association of TIR and cardiovascular autonomic neuropathy (CAN) (12, 17) and in one study, the association of TIR with peripheral nerve function was evaluated (14). Three studies used Medtronic CGM (14, 15, 17), two studies used Freestyle libre (20, 21) while Meiqi (13), and both Meiqi and Medtronic (12) was used in one study each. The CGM data were collected for 3 days in three studies (12-14), for 2 weeks in two studies (20, 21), for both 3 days (GOLD™) and 6 days (iPro^TM2^) in one study (17), and for two 6-day periods, separated by 2 weeks in one study (15).

Each studies evaluating DPN/CAN used different surrogates for examining neuropathy. Three studies used sudomotor function (13), Michigan Neuropathy Screening Instrument scale (15), and Numerical Rating Scale (20) to assess DPN. Kuroda et al. used diagnostic criteria of the Japanese Study Group of Diabetes Neuropathy (22) to diagnose DPN (21). CAN was evaluated in two studies using a combination of five (3-parasympathetic and 2-sympathetic) and four (3-parasympathetic and 1-sympathetic) cardiovascular autonomic function tests (12, 17). Li et al. examined peripheral nerve function by electrophysiologic measurement of motor and sensory nerves to calculate composite Z-score for conduction velocity, latency, and amplitude to assess peripheral nerve function (14).

The prevalence of DPN and CAN was 46.6% and 32.1%, respectively. Yang et al. showed a decline in TIR to be directly associated with increased prevalence of any painful DPN (OR= 2.66, 95 % CI: 1.16–6.10, p < 0.05) (20) while Guo et al. found an increase in TIR to be inversely related to prevalence of sudomotor dysfunction (OR= 0.979, 95% CI: 0.971-0.987, P < 0.001) (13). In one study by Mayeda et al., a 10% decrease in TIR was associated with increase in prevalence of DPN (OR= 1.25, 95% CI: 1.02 - 1.52, p <0.05) and that the rate of DPN was lower in participants with TIR >70 % compared to participants with TIR <70 % (43 % vs. 74%) (15). Kuroda et al. found TIR to be weakly associated with presence of DPN (β = −0.106, P = 0.033) (21). Kim et al. showed the OR of CAN per 10% increase in TIR to be 0.894 (95% CI: 0.81–0.99, p <0.05) (17). Guo et al. found an increase in TIR quartiles to be inversely associated with the prevalence of CAN (P<0.05) (12). Li et al. assessed peripheral nerve function and found higher TIR to be associated with a higher composite Z-score of conduction velocity (b = 0.230, P < 0.001), higher composite Z-score of amplitude (b = 0.099, P = 0.010), and lower composite Z-score of latency (b = 0.172, P < 0.001). The authors further concluded higher TIR tertile group to have lower risk of slowing conduction velocity (TIR medium: OR 0.44, P < 0.001; TIR high: OR 0.26, P < 0.001), lower risk of amplitude reduction (TIR high: OR 0.60, P < 0.05), and higher rate of reduced latency (TIR medium: OR 1.57, P < 0.05; TIR high OR 1.71, P < 0.05) compared to low tertile group (14).

Out of five studies evaluating DPN or peripheral nerve function, three studies evaluated the relationship between TIR and A1c (13-15) while it was not examined in two studies (20, 21). There was no statistically significant between A1c among patients with and without sudomotor dysfunction. One study found higher A1c in patients with sudomotor dysfunction compared to patients without sudomotor dysfunction, however this association was not statistically significant (8.94 vs 8.6, p =0.118) (13). In another study, authors did not find statistically significant relationship between A1c and severity of DPN (p = 0.139) (15). Assessment of nerve function by Li et al. demonstrated A1c to be independently associated with composite Z-score of conduction velocity, latency and amplitude (p < 0.001) (14). Compared to the relationship between A1c and DPN, association between A1c and CAN was limited with only two studies examining this relationship. While one study showed higher A1c to be associated with greater prevalence of CAN (p=0.041) (17) in another study this association trended towards statistical significance (p=0.053)(12).

In summary, increase in TIR was associated with decrease in prevalence and severity of both DPN and CAN. TIR >70 % was associated with significantly lower prevalence of DPN compared to TIR<70%. Additionally, based on limited studies, TIR was found to more closely correlate with DPN and CAN compared to A1c.

## DISCUSSION

To our knowledge, this is the first systematic review examining the relationship between CGM-derived TIR and microvascular complication among patients with T2DM. The risk of microvascular complications and chronic hyperglycemia, as measured by A1c has been well established in both T1DM and T2DM patients in landmark trials such as DCCT and U.K. Prospective Diabetes Study (23, 24). CGM provides more comprehensive glucose data beyond A1c, is convenient for the patients (25), and has constantly shown promising evidence supporting improved glycemic control and quality of life among patients with diabetes (26). Several studies have assessed the relationship between CGM-derived metrics and A1c. One study by Vigersky and McMahon analyzing 18 randomized controlled trials found a strong correlation between A1c and TIR 70–180 mg/dL (27), while Beck et al. found that a 10% change in the TIR 70–180 mg/dL correlates with the mean A1c change by ∼0.5% (28). The current international consensus recommends TIR of 70% to align with A1c of ∼7% with each 10% increase in TIR to correspond with ∼0.5% A1c reduction (4, 5). In a previous work, Beck et al. validated strong correlation between TIR-derived from SMBG among patients in DCCT with the risk of microvascular complications (6). With this association between SMBG-derived TIR and microvascular complications, along with recent data validating the association of CGM-derived TIR with A1c, there is a growing interest in using CGM-derived TIR as a surrogate marker for the assessment of various diabetes-related complications. This idea has been supported in many recent studies and the evidence is constantly growing. One study found a 6.4 % reduction in risk of abnormal carotid intima-media thickness, a surrogate for cardiovascular disease when TIR increased by 10% (29). In a study by Ranjan et al. on patients with T1DM, improved TIR over one year was associated with reduced albuminuria (19% reduction in UACR per 10% increase in TIR) (30). Lu et al. found an inverse correlation between TIR quartile and severity of diabetic retinopathy (11). Yoo et al. reported albuminuria, a microvascular complication of diabetes, to be inversely related with TIR (16) while Myaeda et al. established an association between DPN among patients with T2DM and chronic kidney disease (15). The results of these studies further strengthen the potential of TIR as a tool for predicting the risk of development and progression of diabetes-related microvascular complications. In this systematic review of 11 original published articles, we found a strong correlation between CGM-derived TIR and microvascular complications.

Although CGM-derived TIR provides important information not captured by A1c, it is still insufficient in providing a complete description of glycemic control. Glycemic variability (measured by mean absolute glucose, standard deviation and coefficient of variation) has also been suggested to be an independent predictor of diabetes related complications (31). It results in oxidative stress (32) and endothelial dysfunction (33) and can lead to cardiovascular as well as microvascular complications (34-36). Glycemic variability was not consistently assessed in the 11 studies included in our systematic review, and hence we did not evaluate its association with microvascular complications in our systematic review.

The association between TIR and microvascular complications was adjusted for A1c. The association was independent of A1c for the severity of neuropathic pain (20), sudomotor dysfunction (13), conduction velocity (14), CAN (12), the severity of DR (11), and (albuminuria) (19). Varghese et al. also observed A1c independent association between TIR and retinopathy as well as nephropathy parameters among hypo and hyper profiles versus TIR profiles (18). In three studies, the association between TIR and CAN (17), albuminuria (16) and diabetic retinopathy (19), was found to be dependent on A1c. Additionally, Mayeda et al. did not observe any association between A1c and DPN (15). Parameters, although non-uniformly defined among 11 studies, were consistently applied to the study subjects.

There are some limitations in our systematic review which needs to be noted. The small number of available studies evaluating the association of CGM-derived TIR and microvascular complications and the quality of the studies included in the review are the most significant limitation of our systematic review. The involved studies were primarily cross-sectional in design, and in the absence of prospective studies, a direct causal relationship cannot be established between TIR and microvascular complications. The majority of studies were conducted in Asia (10 out of 11 studies), and study participants were predominantly Asian (13882 out 13987), affecting the generalizability of our findings to other populations. There were significant methodological differences among various studies included in our systematic review. The studies differed in the type of CGM device used (Medtronic vs. Freestyle libre vs. Meiqi), duration over which CGM data were collected (72 hours to 14 days), and calibration of CGM devices (2 to 4 times per day). Additionally, there was no study involving the Dexcom CGM device, one of the most used CGM devices, owing to its ability to communicate with closed-loop systems. These differences could significantly impact the TIR (37). Furthermore, each study used different surrogates for the assessment of neuropathy, retinopathy, and nephropathy. For DR, participants were classified into mild non-proliferative DR, moderate non-proliferative DR, and vision-threatening DR by Lu et al. (11) versus simple DR, pre-proliferative DR, and proliferative DR by Wakasugi et al. (19) versus non-proliferative DR and proliferative DR by Varghese et al. (18). To evaluate DN, patients were categorized into micro and macroalbuminuria in one study versus microalbuminuria, diabetic kidney disease, and macroalbuminuria by Varghese et al. vs. with and without albuminuria in the third study. Similarly, for defining DPN, different criteria like sudomotor dysfunction, Michigan Neuropathy Screening Instrument scale and Numerical Rating Scale were used in three different studies. To evaluate for CAN, one study used 5 cardiac reflex tests while only 4 cardiac reflex tests were used in another study. This heterogeneity in outcomes assessed prevented quantitative analysis in our systematic review. Moreover, only in a few studies (6 out of 11), authors evaluated the association of microvascular complications to a 10 % change in TIR. In contrast, the other studies did not report this data and only assessed the outcomes for different TIR quartiles.

## CONCLUSION

In summary, our study affirms the significant association between CGM-derived TIR and microvascular complications of DN, DR, and DPN among patients with T2DM. However, heterogeneity in the CGM data reported a lack of uniformity in methodology and outcome measured, limited race/ethnicity of the population evaluated in the included studies, and restricted generalization of the findings from our systematic review. Therefore, a largerLscale multi-center investigation that includes more diverse participants is warranted to further validate the association between CGM-derived TIR and microvascular complications among patients with T2DM.

## Supporting information

Not applicable

## Data Availability

All datasets for this study are included in the manuscript and the supplementary files.

## ABBREVIATIONS

TIR: time in range
CGM: continuous glucose monitoring
SMBG: self-monitored blood glucose
DR: diabetic retinopathy
DN: diabetic nephropathy
DPN: diabetic peripheral neuropathy (DPN)
CAN: cardiac autonomic neuropathy
FLP-CGM: Freestyle libre CGM
UACR: urinary albumin-to-creatinine ratio
OR: odds ratio
T1DM: Type 1 diabetes mellitus
T2DM: type 2 diabetes mellitus

## DATA AVAILABILITY STATEMENT

All datasets for this study are included in the manuscript and the supplementary files.

## AUTHOR’S CONTRIBUTION

**Conception and design:** RR

**Development of methodology:** RR, RM

**Data Collection:** RR, RM, NJ

**Analysis and interpretation of data:** RR, RM, NJ, VJ

**Writing, review, and/or revision of the manuscript:** RR, RM, NJ, VJ, RC, PK

**Study supervision:** RR, RC, PK

