## Supplemental File 1 for "Time In Range, as measured by continuous glucose monitor, as a predictor of microvascular complications in type 2 diabetes– A systemic review"

**Supplementary Appendix**

**Contents**

Appendix 1. Details of search strategy

Appendix 2. Characteristics of the studies investigated

Appendix 3. Study quality assessment

**Appendix 1. Details of Search strategy**

| **DATABASE** | **TIMESPAN** | **SEARCH STRATEGY** |
| --- | --- | --- |
| Pubmed | 2010 to June 2021 | (("Continuous glucose monitor" OR "Continuous glucose monitoring" OR "Dexcom" "Freestyle Libre" OR "Guardian" OR "Flash glucose monitoring" OR "Time in Range" OR "Time-in-range" OR "TIR") AND ("Diabetes complications" OR "Microvascular complications" OR "Retinopathy" "Neuropathy" OR "Nephropathy" OR "Complication" OR "Microalbuminuria" OR "Albuminuria")) AND ("Type II diabetes" OR "type 2 diabetes") |
| Scopus | 2010 to June 2021 | (("Continuous glucose monitor" OR "Continuous glucose monitoring" OR "Dexcom" "Freestyle Libre" OR "Guardian" OR "Flash glucose monitoring" OR "Time in Range" OR "Time-in-range" OR "TIR") AND ("Diabetes complications" OR "Microvascular complications" OR "Retinopathy" "Neuropathy" OR "Nephropathy" OR "Complication" OR "Microalbuminuria" OR "Albuminuria")) AND ("Type II diabetes" OR "type 2 diabetes") |
| Web of Science | 2010 to June 2021 | (("Continuous glucose monitor" OR "Continuous glucose monitoring" OR "Dexcom" "Freestyle Libre" OR "Guardian" OR "Flash glucose monitoring" OR "Time in Range" OR "Time-in-range" OR "TIR") AND ("Diabetes complications" OR "Microvascular complications" OR "Retinopathy" "Neuropathy" OR "Nephropathy" OR "Complication" OR "Microalbuminuria" OR "Albuminuria")) AND ("Type II diabetes" OR "type 2 diabetes") |

**Appendix 2. Characteristics of the Studies Investigated**

| **Study ID** | Varghese et al. (Varghese JS, Ho JC, Anjana RM, Pradeepa R, Patel SA, Jebarani S, Baskar V, Narayan KMV, Mohan V. Profiles of Intraday Glucose in Type 2 Diabetes and Their Association with Complications: An Analysis of Continuous Glucose Monitoring Data. Diabetes Technol Ther. 2021 Aug;23(8):555-564. doi: 10.1089/dia.2020.0672. Epub 2021 May 11. PMID: 33720761) |
| --- | --- |
| **Country/Year** | India/2021 |
| **Study Design** | Cross Sectional; Single Centre |
| **Aim Of Study** | To identify profiles of type 2 diabetes from continuous glucose monitoring (CGM) data using ambulatory glucose profile (AGP) indicators and examine the association with prevalent complications. |
| **Methods** | PARTICIPANTS:   - Sample Size: 5901 - SEX (Male % / Female %): 64.8%/ 35.2% - AGE (mean): 55.4 years - ETHNICITY : Asians ( Indian) - DURATION OF DIABETES (mean (SD) years): 8.1(6.8) - Duration during which subjects were recruited:4 years - HbA1c at baseline: 9   INCLUSION CRITERIA:   - age between 18 and 80 years, having at least 75% of CGM data recorded over 14-day period, with valid HbA1c measurements within 30 days of CGM.   EXCLUSION CRITERIA:   - participants with other forms of diabetes |
| **Interventions** | - CGM Devices Used: Abbott FreeStyle Libre Pro- CGM data for two weeks was collected - Duration of CGM use:14 days - Calibration of CGM: NA - % of time CGM device active OR Time wearing CGM :>99% (during 14 days data) |
| **Outcomes/ Result** | - Participants were categorised into three groups based on glycemic profile (‘‘TIR Profile’’, ‘‘Hypo Profile’’, and ‘‘Hyper Profile’’) - Urinary albumin-to-creatinine ratio (UACR) > 30 was used to define DN. Participants were categorized into normoalbuminuria (UACR <30 mg/g), microalbuminuria (UACR 30-300 mg/g) or macroalbuminuria (UACR >300 mg/g). Retinopathy was classified as NPDR and PDR. - Among the participants, 30.1% had retinopathy and 35.5% had nephropathy. - Mean TIR% = 78.4   ASSOCIATION OF CGM PROFILES WITH EXISTING COMPLICATIONS   - Both Hypo and Hyper profiles had higher odds of nonproliferative diabetic retinopathy (“Hypo”: 1.44, 1.20–1.73; “Hyper”: 1.33, 1.11–1.58), macroalbuminuria (“Hypo”: 1.58, 1.25–1.98; “Hyper”: 1.37, 1.10–1.71), and diabetic kidney disease ( “Hypo”: 1.65, 1.18–2.31; “Hyper”: 1.88, 1.37–2.58), compared with “TIR profile. Also, higher odds of PDR (‘‘Hypo profile’’: 2.84, 1.65–4.88 and ‘‘Hyper profile’’: 1.39, 0.78–2.45) as well as NPDR (‘‘Hypo profile’’:1.44, 1.20-1.73 and ‘‘Hyper profile’’:1.33, 1.11-1.58) compared to ‘‘TIR profile’. - The study observed an A1C independent association between TIR and retinopathy and as well as nephropathy parameters among hypo and hyper profiles versus TIR profiles. |
| **Publication details** | COMMERCIAL FUNDING: No  NON-COMMERCIAL FUNDING: No. |

| **Study ID** | Wakasugi et al. (Wakasugi S, Mita T, Katakami N, Okada Y, Yoshii H, Osonoi T, Nishida K, Shiraiwa T, Torimoto K, Kurozumi A, Gosho M, Shimomura I, Watada H. Associations between continuous glucose monitoring-derived metrics and diabetic retinopathy and albuminuria in patients with type 2 diabetes. BMJ Open Diabetes Res Care. 2021 Apr;9(1):e001923. doi: 10.1136/bmjdrc-2020-001923. PMID: 33879513; PMCID: PMC8061826) |
| --- | --- |
| **Country/Year** | Japan/2021 |
| **Study Design** | Exploratory cross-sectional analysis of an ongoing 5year follow up prospective study; Multicentric in 34 institutions |
| **Aim Of Study** | To investigate the relationships between glucose fluctuations evaluated with CGM and the incidence of composite cardiovascular events over a 5-year follow-up period |
| **Methods** | - Sample Size: 999 - AGE (mean (SD) years): 64.6 (9.6) - ETHNICITY : Asians (Japanese) - DURATION OF DIABETES (mean (SD) years): 28.6 (10.8). - Duration during which subjects were recruited: 11 months - HbA1c at baseline: 7   INCLUSION CRITERIA:   - age ≥30 years and ≤80 years, regardless of gender; - receiving treatment for type 2 diabetes at one of the participating outpatient clinics; - no changes (including new prescriptions) in antidiabetic medications for 6 months prior (insulin dosage changes were allowed); - No anticipated changes in antidiabetic medications from the time of enrolment until a CGM device was applied on the back of the upper arm   EXCLUSION CRITERIA:   - type 1 or secondary diabetes - presence of severe infectious disease preoperatively, postoperatively, or associated with severe trauma; - history of myocardial infarction, angina pectoris, cerebral stroke, cerebral infarction, or arteriosclerosis obliterans; current treatment with artificial dialysis; moderate liver dysfunction defined as aspartate aminotransferase ≥100IU/L; moderate or severe heart failure (New York Heart Association stage III or worse); pregnancy, lactation, possible pregnancy, or plans to become pregnant during the study period; present or history of a malignant tumor; - use of a sensor-augmented insulin pump; - type 2 diabetes diagnosis within the past year; |
| **Interventions** | - CGM Devices Used: FreeStyle Libre Pro - Duration of CGM use:14 days; analyzed FLP-CGM data over the middle 8-day period, excluding the first 2 days and last 4 days. - Calibration of CGM: NA - % of time CGM device active OR Time wearing CGM :Analysis of CGM data was done on middle 8-day period during 14 days use |
| **Outcomes/**  **Results** | - The presence and severity of DR were determined by trained ophthalmologists and grouped into four: no diabetic retinopathy (NDR), simple diabetic retinopathy (SDR), pre-proliferative diabetic retinopathy (PPDR), or proliferative diabetic retinopathy (PDR). - DN was defined according to the level of UAE: <30mg/g creatinine was defined as normoalbuminuria, 30–299mg/g creatinine was defined as microalbuminuria, and ≥300mg/g creatinine was defined as macroalbuminuria. - The prevalence of microalbuminuria and macroalbuminuria was 20.3% and 6.7%, respectively. The overall prevalence of diabetic retinopathy was 22.2%; Prevalence of SDR was 13.3%, PPDR 5% and PDR was 3.9%. - Mean TIR% = 78.90   RELATIONSHIP BETWEEN FLP-CGM-DERIVED METRICS AND DR SEVERITY   - Statistically significant association between a 10 % increase in TIR and reduction in severity of DR (OR= 0.85, 95% CI 0.78-0.93, p <0.001) - No significant associations between FLP-CGM-derived metrics and DR severity after adjusting for HbA1c.   RELATIONSHIP BETWEEN FLP-CGM-DERIVED METRICS AND ALBUMINURIA SEVERITY   - Showed statistically significant association between a 10 % increase in TIR and reduction in severity of albuminuria (OR=0.81, 95% CI 0.72- 0.90, p<0.001).The association was independent of A1c for albuminuria. |
| **Publication details** | FUNDING: This study was financially supported by the Japan Agency for Medical Research and Development (AMED) under Grant Number JP20ek0210105 (to HW) and the Manpei Suzuki Diabetes Foundation (to HW).  TO and HW have received research funds from Abbott Japan. HW is a member of the advisory board of Abbott Japan. |

| **Study ID** | Kim et al. (Kim MY, Kim G, Park JY, Choi MS, Jun JE, Lee YB, Jin SM, Hur KY, Kim JH. The Association Between Continuous Glucose Monitoring-Derived Metrics and Cardiovascular Autonomic Neuropathy in Outpatients with Type 2 Diabetes. Diabetes Technol Ther. 2021 Jun;23(6):434-442. doi: 10.1089/dia.2020.0599. Epub 2021 Apr 5. PMID: 33523771) |
| --- | --- |
| **Country/Year** | South Korea/2021 |
| **Study Design** | Cross Sectional Study; Single Center |
| **Aim of Study** | Associations between CGM-derived TIR, hyperglycemia, and hypoglycemia metrics and cardiovascular autonomic neuropathy (CAN) in patients with type 2 diabetes |
| **Methods** | - Sample: 284 - SEX (Male % / Female %): 58.5/ 41.5 - AGE (mean (SD) years): 57.4(10.5) - ETHNICITY : Asians (Korean) - DURATION OF DIABETES (mean (SD) years): 12(8.5) - Duration during which subjects were recruited: 10 years - HbA1c at baseline: 8.30   INCLUSION CRITERIA:   - Those who underwent CGM and autonomic function testing at the same time or within 3 months   EXCLUSION CRITERIA:   - Patients with type 1 or gestational diabetes mellitus - Cancer - History of thyroid dysfunction, myocardial infarction, revascularization, stroke, severe liver disease , GFR <30 mL/min/1.73 m2 |
| **Interventions** | - CGM Devices Used: CGM using GOLD™ (Medtronic MiniMed) for 3 days or iPro™2 (Medtronic MiniMed) for 6 day - Duration of CGM use: 3-days for GOLDTM(Medtronic MiniMed, North Ridge, CA) and 6-days for iProTM2 (Medtronic MiniMed) - Calibration of CGM: 2 - % of time CGM device active OR Time wearing CGM :3 days(GOLD™) or 6 days (iPro™2) (T=77±23.7h; C=76.9±19.2) |
| **Outcomes/ Results** | - Patient were classified in to group with and without cardiovascular autonomic function testing: the cardiovascular autonomic neuropathy (CAN) group (n = 84, 29.6%) and the non-CAN group - Mean TIR %=CAN 57±27; No CAN 62.7 ± 26.8   ASSOCIATIONS BETWEEN TIR 70–180 MG/DL AND CAN   - CAN was evaluated using a combination of five (3-parasympathetic and 2-sympathetic) cardiovascular autonomic function tests. CAN -defined as abnormal result in ≥2 parasympathetic test and severity estimated as sum of scores of 5 Cardiac autonomic function Tests - OR of presence of CAN was 0.876 [95% confidence interval (CI): 0.79–0.98] per 10% increase in the TIR 70–180 mg/dL after adjusting for age, sex, diabetes duration, any medications, and glycemic variability. There is inverse association of severity of CAN with 10% increase in TIR (OR: 0.89, 95% CI: 0.81–0.98) |
| **Publication details** | FUNDING:  This research was supported by a grant of the Korea Health Technology R&D Project through the Korea Health Industry Development Institute (KHIDI), funded by the Ministry of Health and Welfare, Republic of Korea (grant number: HI19C0543). This research was funded by the Korea Disease Control and Prevention Agency (grant number 2020-ER6402-00). |

| **Study ID** | Kuroda et al. (Kuroda N, Kusunoki Y, Osugi K, Ohigashi M, Azuma D, Ikeda H, Makino S, Otsuka A, Tamada D, Watanabe N, Washio K, Tsunoda T, Matsuo T, Konishi K, Katsuno T, Koyama H; Hyogo Diabetes Hypoglycemia Cognition Complications (HDHCC) study group. Relationships between time in range, glycemic variability including hypoglycemia and types of diabetes therapy in Japanese patients with type 2 diabetes mellitus: Hyogo Diabetes Hypoglycemia Cognition Complications study. J Diabetes Investig. 2021 Feb;12(2):244-253. doi: 10.1111/jdi.13336. Epub 2020 Aug 2. PMID: 32594655; PMCID: PMC7858127) |
| --- | --- |
| **Country/Year** | Japan/2021 |
| **Study Design** | Multicentre, Prospective Cohort Study |
| **Aim Of Study** | To assess the relationships between TIR, glycemic variability and patient characteristics in patients with type 2 diabetes mellitus |
| **Methods** | - Sample: 300 - SEX (Male % / Female %): 61.9/ 38.1 - AGE (mean (SD) years): 68 Median, IQR (62-71) - ETHNICITY : Asians (Japanese) - DURATION OF DIABETES (mean (SD) years): 13 Median; IQR 7-23 - HbA1c at baseline: 6.90 - Duration during which subjects were recruited: 2 years   INCLUSION CRITERIA:   - Included patients with type 2 diabetes mellitus, aged between 40 and 75 years, who regularly visited outpatient hospitals or clinics.   EXCLUSION CRITERIA:   - Type 1 diabetes; - Dementia; - Severe hepatic and/or renal dysfunction; - Cancer |
| **Interventions** | - CGM Devices Used: FreeStyle Libre Pro - Duration of CGM use: 10 d (>70% over 14days) - Calibration of CGM: N/A - % of time CGM device active OR Time wearing CGM : 10 d (>70% over 14days) - Mean TIR % = 78.9 (66.9–90.4) |
| **Outcomes/ Results** | ASSOCIATION BETWEEN TIR AND RETINOPATHY   - No statistically significant association between TIR and DR (β = 0.091, P = 0.086)   ASSOCIATION BETWEEN TIR AND NEPHROPATHY   - UACR and EGFR was used - Association was seen between TIR and UACR (β = −0.100, P = 0.043) but not with eGFR (β = −0.011, P = 0.824).   ASSOCIATION BETWEEN TIR AND NEUROPATHY   - Diagnostic criteria of the Japanese Study Group of Diabetes Neuropathy was used to diagnose DPN. - Presence of DPN to be associated with TIR (β = −0.106, P = 0.033) |
| **Publication details** | FUNDING:  This research was funded by the faculty research grant of Hyogo College of Medicine (No. 210790). The authors of the present study thank Drs Hiroyuki Konya (Ashiya Municipal Hospital), Hideki Ifuku (Amagasaki Chuo Hospital), Takeshi Fukui (Fukui Clinic), Isao Hayashi (Hayashi Clinic), Satoru Katayama (Hyogo College of Medicine, Sasayama Medical Center), Masataka Kanyama, Masaru Usami (Ikeda Hospital), Tadahiro Inagaki (Inagaki Medical Clinic), Tomoya Hamaguchi, Chikako Inoue (Itami City Hospital), Akinori Kanzaki (Kawasaki Hospital), Shogo Kurebayashi (Kurebayashi Clinic), Kenji Kusunoki (Kusunoki Clinic), Minoru Kubota (Kwansei Gakuin University, Health Care Center), Takeharu Sasaki (Nishinomiya Watanabe Hospital), Mariko Naka, Sachie Hirose (Osaka Gyoumeikan Hospital), Mitsuyoshi Namba (Takarazuka City Hospital), Tetsuhiro Kitamura (Tamada Clinic) and Hidenori Taniguchi (Taniguchi Medical Clinic). The authors of this study also thank the patients who participated in this study. |

| **Study ID** | Guo et al. (Guo QY, Lu B, Guo ZH, Feng ZQ, Yuan YY, Jin XG, Zang P, Gu P, Shao JQ. Continuous glucose monitoring defined time-in-range is associated with sudomotor dysfunction in type 2 diabetes. World J Diabetes. 2020 Nov 15;11(11):489-500. doi: 10.4239/wjd.v11.i11.489. PMID: 33269061; PMCID: PMC7672791) |
| --- | --- |
| **Country/Year** | China/2020 |
| **Design** | Cross Sectional Study; Single Center |
| **Aim of Study** | To explore the relationship between TIR obtained from CGM and sudomotor function detected by SUDOSCAN in subjects with type 2 diabetes. |
| **Method** | - Sample size: 466 inpatients with T2DM. - SEX (Male % / Female %): 69.9/ 30.1 - AGE (mean (SD) years): 54.5(8.76) - ETHNICITY : Asians (Chinese) - DURATION OF DIABETES (mean (SD) years): 8 Median (range 3-13) - HbA1c at baseline: 8.70 - Duration during which subjects were recruited: October 2017 to May 2019.   INCLUSION CRITERIA:   - Diagnosed according to the 1999 WHO diagnostic criteria for diabetes   EXCLUSION CRITERIA:   - Severe illness or acute stress such as heart failure, liver failure, acute or chronic inflammatory disorders, malignant diseases, and surgery; - History of using oral medications that may affect the nervous system and a recent history of alcoholism; and - Subjects with other metabolic disorders like the lack of vitamin B12. |
| **Interventions** | - CGM Devices Used: CGM (Meiqi Corporation) - Duration of CGM use: 3 days - Calibration of CGM: 4 times - % of time CGM device active OR Time wearing CGM : 3 days - Sudoscan assessment was done using HESC and FESC (sudomotor dysfunction (+) = average FESC <60 µS). |
| **Outcomes/ Result** | - Mean TIR % =SuD+ 72.82 (53.06, 86.79); SuD- 53.12 (28.36, 79.69)   ASSOCITATION BETWEEN TIR and SUDOMOTOR FUNCTION   - Increase in TIR to be inversely related to prevalence of sudomotor dysfunction (OR 0.979, 95% CI: 0.971-0.987, P < 0.001). - The association was independent of A1c. |
| **Publication details** | FUNDING:  By National Natural Science Foundation of China, No.81774134 and No. 81873174; Natural Science Foundation of Jiangsu Province of China, No.BK20150558 and No. BK20171331; Postdoctoral Foundation of Jiangsu Province of China, No. 1501120C; Jiangsu Province 333 Talent Funding Project, No. BRA2017595; and Young Medical Key Talents Project of Jiangsu Province, No.QNRC2016902. |

| **Study ID** | Yoo et al. (Yoo JH, Choi MS, Ahn J, Park SW, Kim Y, Hur KY, Jin SM, Kim G, Kim JH. Association Between Continuous Glucose Monitoring-Derived Time in Range, Other Core Metrics, and Albuminuria in Type 2 Diabetes. Diabetes Technol Ther. 2020 Oct;22(10):768-776. doi: 10.1089/dia.2019.0499. Epub 2020 Apr 13. PMID: 32167394) |
| --- | --- |
| **Country/Year** | South Korea/2021 |
| **Study Design** | Cross Sectional Study; Multicentric; 2 Centres |
| **Aim of Study** | To assess the relationship between TIR and other CGM parameters and the risk of albuminuria in type 2 diabetes |
| **Methods** | - Sample size: 866 - SEX (Male % / Female %): 67.3/ 32.7 - AGE (mean (SD) years): 53 (46-60) - ETHNICITY : Asians( Korean) - DURATION OF DIABETES (mean (SD) years): 8 median (range 3-14) - Duration during which subjects were recruited: 10 years - HbA1c at baseline: 8.20   INCLUSION CRITERIA:   - type 2 diabetes patients   EXCLUSION CRITERIA:   - Subjects with missing data for urine albumin-to creatinine ratio (ACR) (n = 182), subjects with malignancy (n = 5) or severe liver disease defined by a Child-Pugh score of greater than 7 (n = 19), or subjects with an estimated glomerular filtration rate (eGFR) of less than 30 ml/min/1.73m2 when calculated using the Chronic Kidney Disease Epidemiology Collaboration (CKD-EPI) formula (n = 26) were excluded. Subjects with other missing data or with a history of diabetic ketoacidosis or hyperosmolar hyperglycaemic syndrome were also excluded. |
| **Interventions** | - Continuous glucose monitoring was done using Medtronic CGM device for 3 days (GOLD^TM^) and 6 days (iPro^TM2^). - Duration of CGM use: 3 or 6 days - Calibration of CGM: 2 times a day - % of time CGM device active OR Time wearing CGM :T (with albuminuria) =79.9±22.3 hours; C (without albuminuria) =77.5±21.7 h |
| **Outcomes/ Result** | - Urinary albumin-to-creatinine ratio (UACR) > 30 was used to define DN. participants were grouped into with-and without-albuminuria. - Mean TIR% = 78.9 - Prevalence of albuminuria was 36.6%.   ASSOCIATION BETWEEN TIR AND ALBUMINURIA   - Subjects who achieved the target of TIR of 70-180 mg/dL ≥ 70%, TAR (>180 mg/dL) < 25%, and TAR (>250 mg/dL) < 5% had a lower prevalence of albuminuria than subjects who did not achieve those targets (all p < 0.001). - Mean amount of CGM data was 77.5 ± 21.7 hours in subjects without albuminuria and 79.9 ± 22.3 hours in subjects with albuminuria, which were comparable (p = 0.128). Subjects with albuminuria were observed to have significantly lower mean TIR of 70-180 mg/dL and higher TAR > 180 mg/dL , TAR > 250 mg/dL than those without albuminuria. |
| **Publication details** | FUNDING: none |

| **Study ID** | Yang et al. (Yang J, Yang X, Zhao D, Wang X, Wei W, Yuan H. Association of time in range, as assessed by continuous glucose monitoring, with painful diabetic polyneuropathy. J Diabetes Investig. 2021 May;12(5):828-836. doi: 10.1111/jdi.13394. Epub 2020 Sep 29. PMID: 32885597; PMCID: PMC8089011) |
| --- | --- |
| **Country/Year** | China/2021 |
| **Study Design** | Cross Sectional study; Single Center |
| **Aim of Study** | To assess association between TIR and the prevalence and degree of painful diabetic neuropathy (PDN) |
| **Methods** | - Sample size 364 - SEX (Male % / Female %): 67.3/ 32.7 - AGE (mean (SD) years): 53 (46-60) - ETHNICITY : Asians ( Chinese) - DURATION OF DIABETES (mean (SD) years): 8 median (range 3-14) - Duration during which subjects were recruited: 10 months - HbA1c at baseline: 7.35   INCLUSION CRITERIA:   - age>18 years with DPN   EXCLUSION CRITERIA:   - other causes of neuropathy, such as osteoarthritis, cervical and lumbar diseases, connective tissue disease, peripheral vascular disease, tumors, herpes zoster infection, abnormal thyroid function, and severe malnutrition or vitamin B12 deficiency; - coexisting major psychiatric disorders; - severe pain from a cause other than DPN; - central nervous system lesions; - pregnancy. |
| **Interventions** | - CGM Devices: Free Style Libre - Duration of CGM use: 14 days - Calibration of CGM: NA - % of time CGM device active OR Time wearing CGM :2 weeks |
| **Outcomes/ Result** | - The Pain measurement was done using 11 step- numerical rating scale(NRS) and patients were classified into pain free (NRS=0), mild pain (1-3), and moderate/severe pain(4-10) groups - Mean TIR %= 78.90 - Prevalence of painful diabetic neuropathy was 51.92%   ASSOCIATION OF TIR WITH PAINFUL DIABETIC NEUROPATHY   - Compared with the pain-free group, the level of TIR decreased significantly in the mild pain and moderate/severe pain groups (P < 0.05). - The prevalence of mild pain and moderate/severe pain decreased with increasing TIR quartiles (all P < 0.05). - Decline in TIR was directly associated with increased prevalence of any painful DPN (OR 2.66, 95 % CI: 1.16–6.10, p < 0.05). - Multiple linear regression analysis showed that TIR was significantly negatively correlated with the numerical rating scale score after adjustment for glycated hemoglobin, glycemic variability indicators and other risk factors (P < 0.05). - Logistic regression analysis showed decreasing level of TIR was significantly associated with increasing risk of any pain and moderate/severe pain (P < 0.05). - The association was independent of A1c |
| **Publication details** | FUNDING: This study was supported by the National Natural Science Foundation of China (81970705); Central Plains Thousand Talents Plan (204200510026); the Overseas Research and Study Program for Talents in Health Science and Technology of Henan Province (2018078, 2018098). |

| **Study ID** | Li et al. (Li F, Zhang Y, Li H, Lu J, Jiang L, Vigersky RA, Zhou J, Wang C, Bao Y, Jia W. TIR generated by continuous glucose monitoring is associated with peripheral nerve function in type 2 diabetes. Diabetes Res Clin Pract. 2020 Aug;166:108289. doi: 10.1016/j.diabres.2020.108289. Epub 2020 Jun 29. PMID: 32615278) |
| --- | --- |
| **Country/Year** | China/2020 |
| **Study Design** | Cross Sectional Study  Single Center |
| **Aim of Study** | To assess “Association between TIR and nerve conduction study parameters.” |
| **Methods** | - PARTICIPANTS: 740 - SEX (Male % / Female %): 50.95/ 49.05 - AGE (mean (SD) years): 60.24(12.81) - ETHNICITY : Asians (Chinese) - DURATION OF DIABETES (mean (SD) years): 10.66(7.49) - Duration during which subjects were recruited; 1 year and 5 months - HbA1c at baseline: 8.55   INCLUSION CRITERIA:   - Patients with T2DM who had undergone screening for diabetic neuropathy with NCS and were monitored by a CGM system at the inpatient department of Shang-hai Jiao Tong University Affiliated Sixth People’s Hospital between April 2013 and August 2014.   EXCLUSION CRITERIA:   - Progressive malignancy, - Diseases affecting nerve conduction function (such as chronic inflammatory demyelinating polyneuropathy or carpal tunnel syndrome, etc.), o - Other life-shortening medical conditions |
| **Interventions** | - CGM Devices Used: Medtronic CGM device readings were obtained for 3 consecutive days. - Duration of CGM use: 3 days. - Calibration of CGM: NA - % of time CGM device active OR Time wearing CGM : NA - Mean TIR % = not described - Electrophysiologic measurement of motor and sensory nerve was used to calculate composite Z-score for conduction velocity (CV), latency, and amplitude to assess peripheral nerve function. - Parameter for motor nerve studies included (median, ulnar and tibial nerves) - Conduction velocity (CV), compound muscle action potential amplitude (CMAP), distal latency. - Parameter for sensory nerve studies included (median, ulnar, and sural nerves) - Onset latency, sensory nerve action potential amplitude (SNAP), and CV. |
| **Outcomes/ Result** | ASSOCIATION BETWEEN TIR AND NERVE FUNCTION   - Higher TIR was associated with a higher composite Z-score of CV (b = 0.230, P < 0.001), higher composite Z-score of amplitude (b = 0.099, P = 0.010), and lower composite Z-score of latency (b = 0.172, P < 0.001). - Higher TIR tertile group was found to have lower risk of slowing conduction velocity (TIR medium: OR 0.44, P < 0.001; TIR high: OR 0.26, P < 0.001), lower risk of amplitude reduction (TIR high: OR 0.60, P < 0.05), and higher rate of reduced latency (TIR medium: OR 1.57, P < 0.05; TIR high OR 1.71, P < 0.05) compared to low tertile group |
| **Publication details** | FUNDING:  This study was financially supported by National Key Research and Development Programme of China (grant no. 2017YFC0906903), the Municipal Human Resources Development Programme for Outstanding Leaders in Medical Disciplines in Shanghai (grant no. 2017BR045), and the National Human Genetic Resources Sharing Service Platform (grant no. YCZYPT [2017]02), Shanghai Major Clinical Disease Clinical Sample Pool of Professional and Technical Services Platform (18DZ2294100). |

| **Study ID** | Guo et al. (Guo Q, Zang P, Xu S, Song W, Zhang Z, Liu C, Guo Z, Chen J, Lu B, Gu P, Shao J. Time in Range, as a Novel Metric of Glycemic Control, Is Reversely Associated with Presence of Diabetic Cardiovascular Autonomic Neuropathy Independent of HbA1c in Chinese Type 2 Diabetes. J Diabetes Res. 2020 Feb 6;2020:5817074. doi: 10.1155/2020/5817074. PMID: 32090120; PMCID: PMC7026737) |
| --- | --- |
| **Country/Year** | China/2020 |
| **Design** | Cross Sectional Study |
| **Aim of Study** | To explore the relationship between TIR obtained from CGM and sudomotor function detected by SUDOSCAN in subjects with type 2 diabetes. |
| **Method** | - Sample size: 466 inpatients with T2DM. - SEX (Male % / Female %): 69.9/ 30.1 - AGE (mean (SD) years): 54.5(8.76) - ETHNICITY : Asians (Chinese) - DURATION OF DIABETES (mean (SD) years): 8 Median (range 3-13) - HbA1c at baseline: 8.70 - Duration during which subjects were recruited: October 2017 to May 2019.   INCLUSION CRITERIA:   - Diagnosed according to the 1999 WHO diagnostic criteria for diabetes   EXCLUSION CRITERIA:   - Severe illness or acute stress such as heart failure, liver failure, acute or chronic inflammatory disorders, malignant diseases, and surgery; - History of using oral medications that may affect the nervous system and a recent history of alcoholism; and - Subjects with other metabolic disorders like the lack of vitamin B12. |
| **Interventions** | - CGM Devices Used: CGM (Meiqi Corporation) - Duration of CGM use: 3 days - Calibration of CGM: 4 times - % of time CGM device active OR Time wearing CGM : 3 days - Sudoscan assessment was done using HESC and FESC (sudomotor dysfunction (+) = average FESC <60 µS). |
| **Outcomes/ Result** | - Mean TIR % =SuD+ 72.82 (53.06, 86.79); SuD- 53.12 (28.36, 79.69)   ASSOCITATION BETWEEN TIR and SUDOMOTOR FUNCTION   - Increase in TIR to be inversely related to prevalence of sudomotor dysfunction (OR 0.979, 95% CI: 0.971-0.987, P < 0.001). - The association was independent of A1c. |
| **Publication details** | FUNDING:  By National Natural Science Foundation of China, No.81774134 and No. 81873174; Natural Science Foundation of Jiangsu Province of China, No.BK20150558 and No. BK20171331; Postdoctoral Foundation of Jiangsu Province of China, No. 1501120C; Jiangsu Province 333 Talent Funding Project, No. BRA2017595; and Young Medical Key Talents Project of Jiangsu Province, No.QNRC2016902. |

| **Study ID** | Mayeda et al. (Mayeda L, Katz R, Ahmad I, Bansal N, Batacchi Z, Hirsch IB, Robinson N, Trence DL, Zelnick L, de Boer IH. Glucose time in range and peripheral neuropathy in type 2 diabetes mellitus and chronic kidney disease. BMJ Open Diabetes Res Care. 2020 Jan;8(1):e000991. doi: 10.1136/bmjdrc-2019-000991. PMID: 31958307; PMCID: PMC7039577) |
| --- | --- |
| **Country** | USA/2020 |
| **Design** | Cross Sectional Study; Multicentric |
| **Aim of Study** | To assess “describe the prevalence of DPN symptoms among participants with type 2 DM and moderate-to severe CKD and 2) examine the association of glycemia (as measured by CGM) with DPN symptoms among our target population” |
| **Methods** | - Sample Size: 105 [81 participants with moderate-to-severe CKD and 24 control participants] - SEX (Male % / Female %): 63.8/ 36.2 - AGE (mean (SD) years): 67.08(10.09) - ETHNICITY : White, Black, Hispanic, Others - DURATION OF DIABETES (mean (SD) years): 19.08(10.19) - Duration during which subjects were recruited: 2 years - HbA1c at baseline: 7.84   INCLUSION CRITERIA:   - Moderate-to-severe CKD (estimated glomerular filtration rate (eGFR) <60mL/ min/1.73m2).   EXCLUSION CRITERIA:   - age <18 years, history of kidney transplant, dialysis treatment, treatment with erythropoietin, current use of clinical CGM, pregnancy or current therapy for cancer and inability to speak English. |
| **Interventions** | - CGM Devices Used: Medtronic - Duration of CGM use: Two 6-day periods, separated by 2 weeks. - Calibration of CGM: 2 times a day - % of time CGM device active OR Time wearing CGM : Two 6-day periods, separated by 2 weeks. - DPN symptoms was evaluated using MNSI (Michigan Neuropathy Screening Instrument) questionnaire, (Positive if MNSI ≥2 Symptoms) |
| **Outcomes/ Result** | - Mean TIR % =78.90 - DPN prevalence was 74%   ASSOCIATION BETWEEN TIR AND DPN   - Rate of DPN was higher in participants with TIR >70 % compared to participants with TIR <70 % (43 % vs 74%). - DPN prevalence was inversely correlated with TIR (OR 1.25 (95% CI 1.02 to 1.52) per 10% lower TIR). - Lower TIR and higher glucose monitoring indicators (GMI) were associated with DPN symptoms. - 10 % increase in TIR was found to be inversely associated with prevalence of DPN (OR 1.25, 95% CI: 1.02 - 1.52, p <0.05). - Authors did not observe any association between A1c and DPN. |
| **Publication details** | FUNDING:  The CANDY Study was primarily supported by American Diabetes Association grant #4-15-CKD-20. Additional funding came from grants R01DK088762, R01DK087726 and T32DK007247 from the National Institute of Diabetes and Digestive and Kidney Diseases; a grant from Puget Sound Veterans Affairs Health Care System and an unrestricted grant from Northwest Kidney Centres. |

| **Study ID** | Lu et al. (Lu J, Ma X, Zhou J, Zhang L, Mo Y, Ying L, Lu W, Zhu W, Bao Y, Vigersky RA, Jia W. Association of Time in Range, as Assessed by Continuous Glucose Monitoring, With Diabetic Retinopathy in Type 2 Diabetes. Diabetes Care. 2018 Nov;41(11):2370-2376. doi: 10.2337/dc18-1131. Epub 2018 Sep 10. PMID: 30201847) |
| --- | --- |
| **Country/Year** | China/2018 |
| **Study Design** | Cross Sectional study; Single Center |
| **Aim of Study** | To assess “Association between TIR assessed by CGM and DR” |
| **Methods** | - Sample size: 3262 - SEX (Male % / Female %): 50.95/ 49.05 - AGE (mean (SD) years): 60.24 (11.2). - ETHNICITY : Asians ( Chinese) - DURATION OF DIABETES (mean (SD) years): 28.6 (12.81). - Duration during which subjects were recruited: 7 years - HbA1c at baseline: 8.90   INCLUSION CRITERIA:   - age 18 years, presence of type 2 diabetes, and a stable glucose-lowering regimen over the previous 3 months.   EXCLUSION CRITERIA:   - diabetic ketoacidosis; a hyperglycemic hyperosmolar state or severe and recurrent hypoglycemic events within the previous 3 months; and a history of malignancy, mental disorders, or severe kidney or liver dysfunction. |
| **Interventions** | - CGM Devices Used: Medtronic CGM device readings over 3 days - Duration of CGM use: 3 days - Calibration of CGM: 4 times a day - % of time CGM device active OR Time wearing CGM :72h - Participants were classified into mild NPDR, moderate NPDR, and vision threatening DR |
| **Outcomes** | - Mean TIR %= 78.90 - Overall prevalence of DR was 23.9%. The prevalence of mild NPDR, moderate NPDR, and VTDR were 10.9%, 6.1%, and 6.9%, respectively.   ASSOCIATION BETWEEN TIR and DR   - Multinomial logistic regression revealed significant associations between TIR and all stages of DR (mild NPDR, P = 0.018; moderate NPDR, P = 0.014; VTDR, P = 0.019) after controlling for age, sex, BMI, diabetes duration, blood pressure, lipid profile, and HbA1c. - All of the patients were stratified according to quartiles of TIR (quartile 1 [Q1]: #51%; quartile 2 [Q2]: 51–71%; quartile 3 [Q3]: 71–86%; quartile 4 [Q4]: .86%). In general, the prevalence of DR by severity decreased with ascending quartiles of TIR (all P for trend ,0.001). the highest TIR quartile was independently associated with all stages of DR, compared with the lowest quartile (mild NPDR: odds ratio [OR] 0.56, P = 0.010; moderate NPDR: OR 0.48, P = 0.009; VTDR: OR 0.53, P = 0.023) - An inverse correlation between TIR quartile and severity of diabetic retinopathy with statistically significant association between a 10 % increase in TIR and reduction in severity of DR was found. |
| **Publication details** | FUNDING: This work was funded by the National Natural Science Foundation of China (grant no.81100590), the Shanghai United Developing Technology Project of Municipal Hospitals (grant nos. SHDC12006101 and SHDC12010115), and the Shanghai Municipal Education Commission Gaofeng Clinical Medicine grant support (grant no. 20161430). |

**Appendix 3. Study Quality Assessment**

|  | **Major Outcomes** | **Varghese et. al. 2021** | **Wakasugi et. al. 2020** | **Kim et. al. 2021** | **Kuroda et. al. 2021** | **Guo et. al. 11/2020** | **Yoo et. al. 2020** | **Yang et. al. 2020** | **Li et. al. 2020** | **Guo et. al. 02/2020** | **Mayeda et. al. 2019** | **Lu et. al. 2018** |
| --- | --- | --- | --- | --- | --- | --- | --- | --- | --- | --- | --- | --- |
| 1 | Was the research question or objective in this paper clearly stated? | Yes | Yes | Yes | Yes | Yes | Yes | Yes | Yes | Yes | Yes | Yes |
| 2 | Was the study population clearly specified and defined? | Yes | Yes | Yes | Yes | Yes | Yes | Yes | Yes | Yes | Yes | Yes |
| 3 | Was the participation rate of eligible persons at least 50%? | Yes | Yes | Yes | Yes | Yes | Yes | Yes | Yes | Yes | Yes | Yes |
| 4 | Were all the subjects selected or recruited from the same or similar populations? Were inclusion and exclusion criteria prespecified and applied uniformly to all participants? | Yes | Yes | Yes | Yes | Yes | Yes | Yes | Yes | Yes | Yes | Yes |
| 5 | Was a sample size justification, power description, or variance and effect estimates provided? | No | No | No | No | No | Yes | No | No | No | No | No |
| 6 | For the analyses in this paper, were the exposure(s) of interest measured prior to the outcome(s) being measured? | No | No | No | No | No | Yes | No | No | No | Yes | No |
| 7 | Was the timeframe sufficient so that one could reasonably expect to see an association between exposure and outcome if it existed? | No | Yes | Yes | Yes | No | Yes | No | No | Yes | Yes | Yes |
| 8 | For exposures that can vary in amount or level, did the study examine different levels of the exposure as related to the outcome ? | Yes | Yes | No | Yes | Yes | Yes | Yes | Yes | Yes | Yes | Yes |
| 9 | Were the exposure measures (independent variables) clearly defined, valid, reliable, and implemented consistently across all study participants? | Yes | Yes | Yes | Yes | Yes | Yes | Yes | Yes | Yes | Yes | Yes |
| 10 | Was the exposure(s) assessed more than once over time? | Yes | Yes | Yes | Yes | Yes | No | Yes | Yes | Yes | Yes | Yes |
| 11 | Were the outcome measures (dependent variables) clearly defined, valid, reliable, and implemented consistently across all study participants? | Yes | Yes | Yes | Yes | Yes | Yes | Yes | Yes | Yes | Yes | Yes |
| 12 | Were the outcome assessors blinded to the exposure status of participants? | No | No | No | N/A | No | Yes | N/A | N/A | No | No | No |
| 13 | Was loss to follow-up after baseline 20% or less? | Yes | Yes | Yes | Yes | Yes | Yes | Yes | Yes | Yes | Yes | Yes |
| 14 | Were key potential confounding variables measured and adjusted statistically for their impact on the relationship between exposure(s) and outcome(s)? | Yes | No | Yes | Yes | Yes | Yes | Yes | Yes | Yes | Yes | Yes |
|  | **Overall Quality Rating** | Fair | Fair | Fair | Fair | Fair | Good | Fair | Fair | Fair | Fair | Fair |
